# A multi-center investigation on the effect of decompressive surgery on Balance and physical ActiviTy Levels in patients with lumbar Spinal stenosis (B-ATLAS): Protocol for a prospective cohort study

**DOI:** 10.1101/2024.02.20.24303085

**Authors:** OB. Zielinski, DW. Hallager, KY. Jensen, LY. Carreon, M.Ø. Andersen, LP. Diederichsen, RD. Bech

## Abstract

**Background:** Patients with lumbar spinal stenosis may complain of poor balance, decreased physical function and problems maintaining physical activity levels due to radiculopathy. Decompressive surgery is often indicated if conservative management fails to achieve a satisfactory clinical outcome. While surgical management has proven effective at treating radiculopathy, and patients report increased physical function post-operatively, objective measures of postural control and physical activity remains sparse. This study aims to investigate the effects of decompressive surgery on balance and activity levels of elderly patients with lumbar spinal stenosis using objective measurements.

**Methods and analysis:** This is a 24-month, multi-centre, prospective cohort study. Patients ≥ 65 years of age with MRI-verified symptomatic lumbar spinal stenosis will be recruited from two separate inclusion centres, and all participants will undergo decompressive surgery for lumbar spinal stenosis. Pre-operative data is collected up to 3 months before surgery, with follow-up data collected at 3, 6, 12 and 24-months post-operatively. Balance measurements are performed using the Wii Balance Board, mini–Balance Evaluation Systems Test and Tandem test, and data concerning physical activity levels are collected using ActiGraph wGT3X-BT accelerometers. Patient reported outcomes regarding quality of life and physical function are collected from the EuroQol-5D, 36-Item Short Form Health Survey and Zurich Claudication Questionnaire. Primary outcomes are the change in sway area of centre of pressure and total activity counts per day from baseline to follow-up at 24-months. A sample size of 80 participants has been calculated.

**Ethics and dissemination:** The study has been approved by the Regional Ethics Committee of Region Zealand (ID EMN-2022-08110) and the Danish Data Protection Agency (ID REG-100-2022). All results from the study will be published in international peer-reviewed journals and presented at national and international conferences. Study findings will be disseminated through national patient associations.

**Trial registration number:** NCT06075862 & NCT06057428

**STRENGTHS AND LIMITATIONS:**

- This study will provide new knowledge concerning the effects of lumbar spinal decompression surgery on postural control and physical activity levels.
- A follow-up period of two years ensures that changes in the primary outcomes are sufficiently documented.
- This will be the first study to correlate changes in postural control with changes in physical activity levels amongst patients with lumbar spinal stenosis.
- A pre-post prospective cohort design provides the necessary comparative control group.

## INTRODUCTION

### Background

Lumbar spinal stenosis (LSS) is one of the most common degenerative diseases of the spinal column, with prevalence rising with age and up to 14% affected amongst patients ≥ 60 [1,2]. Although often asymptomatic, common symptoms of LSS include low back pain as well as lumbar radiculopathy that worsens with prolonged ambulation, lumbar extension, and standing [3-5]. Patients may also complain of poor balance, decreased physical function and problems maintaining physical activity levels, and examination findings may include a wide-based gait and abnormal Romberg results [5-7]. LSS has been shown to be a risk factor for both postural imbalance as well as falls, and patients with LSS are often physically inactive due to the ambulatory limitations that symptomatic LSS can present with, despite evidence suggesting the benefits of physical activity [8-14].

Non-operative management has been suggested as first-line treatment for LSS, with a combined approach of physical therapy and pharmacological treatment with NSAIDs and analgesics, though surgical decompression is often the treatment of choice in patients with ongoing pain despite conservative management for 3-6 months [5,15-20]. The effect of surgical decompression on disability and leg pain has been widely evaluated, and patients report overall satisfactory outcomes. However, studies of the effect on postural control and physical activity levels are scarce [17]. While patients report subjective improvements in physical activity and balance following surgery, instrumented objective measurements are lacking, and correlations with patient reported quality-of-life and physical function are not well understood [20,21]. This study aims to rectify this by investigating the postural control and physical activity of patients with LSS before and after decompressive surgery using a combination of validated objective instruments and patient-reported outcome measures.

### Postural control

Any deficit relating to somatosensory, motor control or cognitive systems associated with postural control can lead to postural imbalance. These deficits can include a wide range of pathological conditions such as visuo-vestibular and neurodegenerative diseases, as well as orthopaedic and rheumatological diseases of the musculoskeletal system [22-24]. Efforts have been made to quantify the degree of postural imbalance in patients with spinal stenosis compared to healthy controls and small inroads have been made in determining the effect of decompressive surgery, though the true effect size remains unclear [9,25-28].

Various methods of assessing postural control have been developed, though as of writing no gold-standard method exists. Static posturography, which aims to measure postural control by quantifying center of pressure (COP) changes during quiet standing, has been highlighted as an objective and highly granular method, with the ability to predict falls amongst older adults [29-31]. Posturography has traditionally been performed using a force plate, capable of measuring ground reaction forces and moments, allowing for the quantification of positional and dynamic variables associated with COP position, displacement, and trajectory [32,33]. The Wii Balance Board (WBB) (Nintendo Co., Ltd., Kyoto, Japan), first released in 2007, has recently gained popularity as a low-cost, portable force-plate transducer, for use in postural control assessment. The WBB has been extensively validated against laboratory-grade force plates and shown to have good intra-device and inter-device reliability, as well as being a valid tool to measure balance amongst older adults [34,35]. Work has been done to ensure that the WBB can be calibrated to minimize measurement error, and open-source algorithms exist for the calculation of positional and dynamic variables recorded by the WBB [33,36]. As such, the use of the WBB in longitudinal monitoring for research purposes is considered viable [37].

### Activity levels

Physical activity has been demonstrated to be correlated to physical and mental wellbeing, offering significant benefits including preventing and managing cardiovascular disease, cancer, and diabetes, as well as reducing symptoms of depression and anxiety [14,38,39]. The negative consequences of sedentary behaviour are likewise well established, increasing risks of metabolic and musculoskeletal disorders as well as all-cause mortality [40]. As such, physical activity as both an intervention tool and as a measurement of effect has been becoming increasingly prevalent in the literature.

The use of accelerometers as an activity monitoring device in intervention studies has become increasingly prevalent due to the high frequency of measurements, large memory capacity, low subject interference and ability to differentiate between differing levels of activity [41]. Accelerometer use has been recommended as a clinical measurement of physical activity when undertaking intervention studies and has seen a rise in use in the field of orthopaedics [42-45].

The ActiGraph wGT3X-BT (ActiGraph, LLC, Ft. Walton Beach, FL, USA) is a triaxial accelerometer, recording inertia in three planes at a sampling rate up to 100 Hz. A proprietary filter can be applied to eliminate artifacts due to movement not caused by human activity, and data is summed as total activity count per minute, which can then be used to estimate physical activity energy expenditure (PAEE) and time spent in moderate-vigorous physical activity (MVPA). The wGT3X-BT has been widely validated against gold standard measurements such as doubly labelled water, and in appropriate patient groups such as the elderly, and has proven valid and reliable in assessing physical activity intensity [46-48].

Studies on physical activity levels after decompressive surgery amongst patients with LSS has so far been unable to demonstrate a significant effect six months post-operatively measured by accelerometer [21]. However, while comparable studies on patients undergoing total hip arthroplasty likewise found no significant improvement in activity levels after six months, studies with longer follow-up, up to a year post-operatively, have been able to show an activity level comparable to healthy control individuals [49,50]. Questions persist however regarding retention of potential increases in physical activity in the years following decompressive surgery.

### Study aims and hypotheses

The primary aim of this study is to investigate and quantify the changes in postural control and activity levels amongst elderly patients with lumbar central canal spinal stenosis following decompressive surgery, between baseline and 24-month follow-up measurements. Secondary aims are to correlate findings concerning postural control and activity levels with patient-reported physical function and health-related quality of life outcomes.

## METHODS AND ANALYSIS

### Study design and setting

This is a 24-month, multi-centre, prospective cohort study conducted at Zealand University Hospital Køge, Denmark and the Spine Centre of Southern Denmark, Middelfart. Participants in this study are patients with symptomatic MRI (Magnetic Resonance Imaging)-verified central canal spinal stenosis undergoing in-patient surgical decompression at either centre. Measurements of postural control and activity will be undertaken up to 3 months before decompressive surgery, as well as 3, 6, 12, and 24 months post-operatively, as presented in Figure 1. A recruitment phase of 12 months is planned, and study end is defined as ‘last patient out’. Study inclusion began in September 2023, and the study is expected to be completed December 2026.

**Figure 1:**
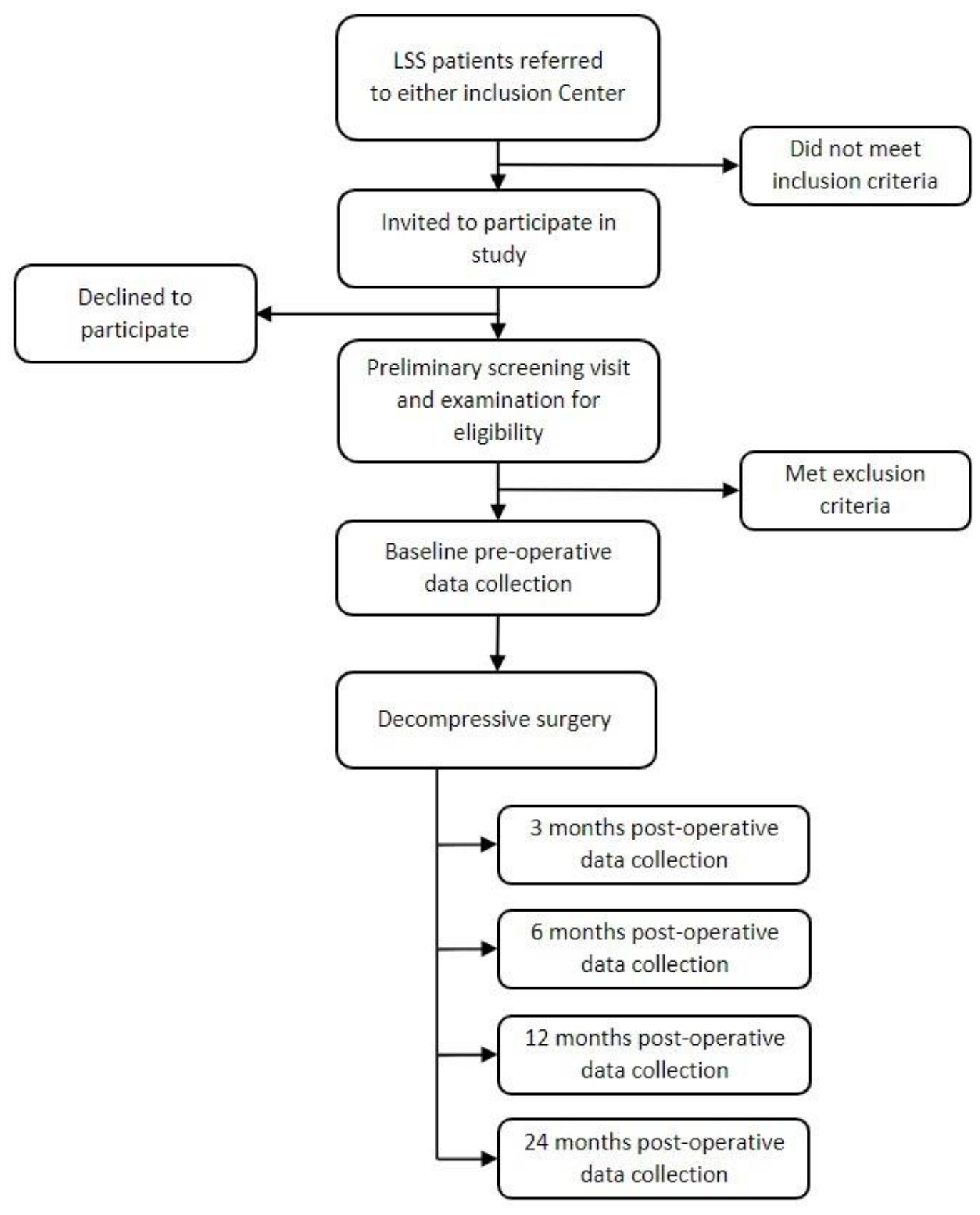
Flowchart of participant timeline and patient recruitment flow during the study period.

### Recruitment

Patients referred for surgical decompression at either the Spine Section at Zealand University Hospital, Køge or the Spine Center of Southern Denmark, Middelfart, will be invited to participate. The full scope of the study, as well as any associated risks and benefits, will be explained prior to the collection of informed consent. Participants will be assessed for eligibility (see below), and eligible participants will undergo pre-operative data collection procedures prior to their scheduled surgery.

### Eligibility criteria

#### Inclusion criteria

(1) age ≥ 65 years at time of evaluation, (2) planned for decompressive spinal surgery due to symptomatic LSS at ≥ 1 level, (3) central canal LSS grade C or D by Schizas’ classification [51] at ≥ 1 level, verified by MRI, and (4) a minimum of 3 months of unsuccessful non-operative treatment.

#### Exclusion criteria

(1) signs of malignancy or infection in the spinal column, (2) severe comorbidities defined as American Society of Anaesthesiologist’s (ASA) physical status classification score ≥ 3, (3) revision surgery defined as previous decompressive surgery at the same vertebral level, (4) any spinal surgery up to one year prior to the date of evaluation, (5) Mini Mental State Examination score ≤ 27, or (6) degenerative spondylolisthesis ≥ 3mm on pre-operative imaging.

Participants may withdraw from the study at any time. Participants will be inquired as to their reason for withdrawal, though providing a reason is not mandatory. Reasons for withdrawal, as well as causes of loss to follow-up, will be recorded.

### Intervention

All patients will undergo intervention in the form of open decompressive surgery at ≥ 1 vertebral level due to central spinal stenosis. Treatment before and after the surgical procedure will remain as standard-of-care, including rehabilitative and physiotherapeutic efforts.

### Outcome measures

#### Primary outcome measure of postural control

The primary outcome measure will be the mean change in sway area of COP (cm^2^) from baseline (pre-operative) to 24 months post-operative. Sway area denotes the area in which COP oscillates during quiet standing and will be defined as the area of the ellipse which contains the true mean of (Y_n_,X_n_)_1≤n≤N_ with 95% probability, as derived by Schubert and Kirchner [52].

Measures of postural control will be calculated from the vertical forces recorded from the WBB (Nintendo Co., Ltd., Kyoto, Japan) using the BrainBLoX interface developed by the neuromechanics laboratory at University of Colorado Boulder [53]. The WBB will be calibrated according to guidelines set forth by Clark et al [34]. As data from the WBB is sampled at a variable rate, a Sliding Window Average with Relevance Interval Interpolation method will be used to resample the signal at 100Hz. The resulting data will be processed using a fourth-order, low-pass Butterworth filter with a 10Hz cutoff frequency and centered using the arithmetic mean. Postural control measures will be calculated using custom software based on standardized methods set forth by Quijoux et al [33].

Consultations will be conducted in an unperturbed room. Measurements are taken while patients stand on the WBB in a standardized and consistent location, with their arms hanging by their sides, and being told to focus on a visible point ahead of them at head height. Patients will be told to stand as still as possible [54].

Measurements are recorded through two sets of five trials of 90 seconds each, with eyes open and eyes closed respectively. Rests of 15 second intervals are given between each trial, and a rest of 60 seconds will be given between each set. The number of trials and the lengths of these are based upon recommendations from reliability studies by Doyle et al and Ruhe et al [54,55]. A Visual Analog Scale (VAS) concerning current degree of leg pain will be recorded before each set.

The need to take a corrective step during the trial to avoid falling will be recorded.

#### Secondary outcome measures of postural control

Secondary outcome measures will be change in mean COP velocity in the antero-posterior and medio-lateral plane (mm) from baseline to 24-months post-operative, and change in the Danish version of the Mini Balance Evaluation Systems Test (Mini-BESTest) and Tandem test score from baseline to 24-months post-operative. The Mini-BESTest was initially developed to aid in the analysis of various postural control systems while being brief and simple to administer; it has since been validated for use in elderly populations and has previously been applied in studies concerning patients with LSS to identify various balance limitations [56-59].

#### Primary outcome measure of activity level

The primary outcome measure will be the mean change in Total Activity Counts per day (TAC/d) from baseline to 24 months post-operative, as measured by the ActiGraph wGT3X-BT accelerometer.

At each data collection visit, participants will be instructed to wear the accelerometer for seven consecutive days and instructed to wear the accelerometer on their hip during all waking hours except when bathing or swimming. A wear-time diary will be issued, and participants will be instructed in their use.

The accelerometer will be set to sample at a rate of 30 Hz, and an epoch length of 60 seconds will be implemented. As the study population concerns elderly patients, the low-frequency extension filter will be applied, to better help capture slower movements that may be prevalent amongst this group. Non-wear time will be defined using the Choi et al algorithm, consisting of 90 minutes of zero counts per minute (cpm) with an allowance of 2 minutes of activity when it is placed between two 30-minute windows of zero cpm [60]. Sampling rate, epoch length, filter choice and non-wear-time definition are chosen in accordance with recommendations set forth by Migueles et al for older adults, while activity intensity levels (sedentary, light, moderate and vigorous intensity) will be defined using cut-points generated and validated by Bammann et al [31,47,38].

To achieve a suitable degree of intraclass correlation between measurement intervals, a valid day of measurement will be defined as awake wear-time ≥ 10 hours per day, and a valid week of measurement will be defined as ≥ 4 valid days of wear-time during the seven-day interval [31].

#### Secondary outcome measures of physical activity

Secondary outcome measures will be change in time spent MVPA per day (min/d) and sedentary time (hours/d) from baseline to 24-months post-operative, as calculated from accelerometer data, and change in self-reported activity level as measured by International Physical Activity Questionnaire short form (IPAQ-SF) score. The IPAQ-SF will be issued to all patients upon completion of a week of activity measurements using the wGT3X-BT accelerometer. While the validity of the IPAQ-SF concerning absolute measurements of activity is dubious, for the purpose of estimating self-reported physical activity, the IPAQ-SF has proven sufficient [61].

#### Patient-reported outcome measures

Primary outcomes of postural control and activity levels will be compared to patient-reported outcome measures of quality-of-life and physical function as additional secondary outcome measures. Change in quality-of-life and physical function will be collected through surveys using the Danish versions of the European Quality of Life – 5 Dimensions (EQ-5D) questionnaire, the Short Form Health Survey (SF-36) and the Zurich Claudication Questionnaire (ZCQ). Both the EQ-5D and SF-36 have been translated and validated for use in Danish and are used extensively in clinical settings in Denmark [62,63]. The ZCQ, also known as the Swiss Spinal Stenosis Measure, was initially developed for the purpose of assessing symptom severity for LSS, physical function, and surgical management satisfaction [64,65].

All data collection tools used in the study will be administered by either the Principal Investigator (PI) or a licensed physiotherapist employed at either study centre, trained in the use of the data collection tools by the PI. For an overview of data collection tools administered during the study period, see table 1. The order of tests during each data collection visit will remain consistent to account for subject fatigue. For an overview of the order of tests, see Figure 2.

**Table 1:** Overview of the data collections tools administered during the course of the study. *\*during the previous 3 months; **questions 13 through 18 pertaining to the effect of surgical treatment are only administered at post-operative data collection visits. WBB: Wii Balance Board*.

| *Data collection tool* | **Baseline, pre-operative** | **3m post-operative** | **6m post-operative** | **12m post-operative** | **24m post-operative** |
| --- | --- | --- | --- | --- | --- |
| ***Electronic Health Record*** |  |  |  |  |  |
| Comorbidities | X |  |  |  |  |
| Spinal history | X |  |  |  |  |
| Medications | X | X | X | X | X |
| ***Participant interview*** |  |  |  |  |  |
| Height | X |  |  |  |  |
| Weight | X |  |  |  |  |
| History of falls* | X | X | X | X | X |
| ***Activity monitoring*** |  |  |  |  |  |
| wGT3X-BT accelerometer | X | X | X | X | X |
| ***Balance measurements*** |  |  |  |  |  |
| WBB, eyes open | X | X | X | X | X |
| WBB, eyes closed | X | X | X | X | X |
| mini-BESTest | X | X | X | X | X |
| Tandem test | X | X | X | X | X |
| ***Patient reported outcome measures*** |  |  |  |  |  |
| EQ-5D | X | X | X | X | X |
| SF36 | X | X | X | X | X |
| ZCQ** | X | X | X | X | X |

**Figure 2:**
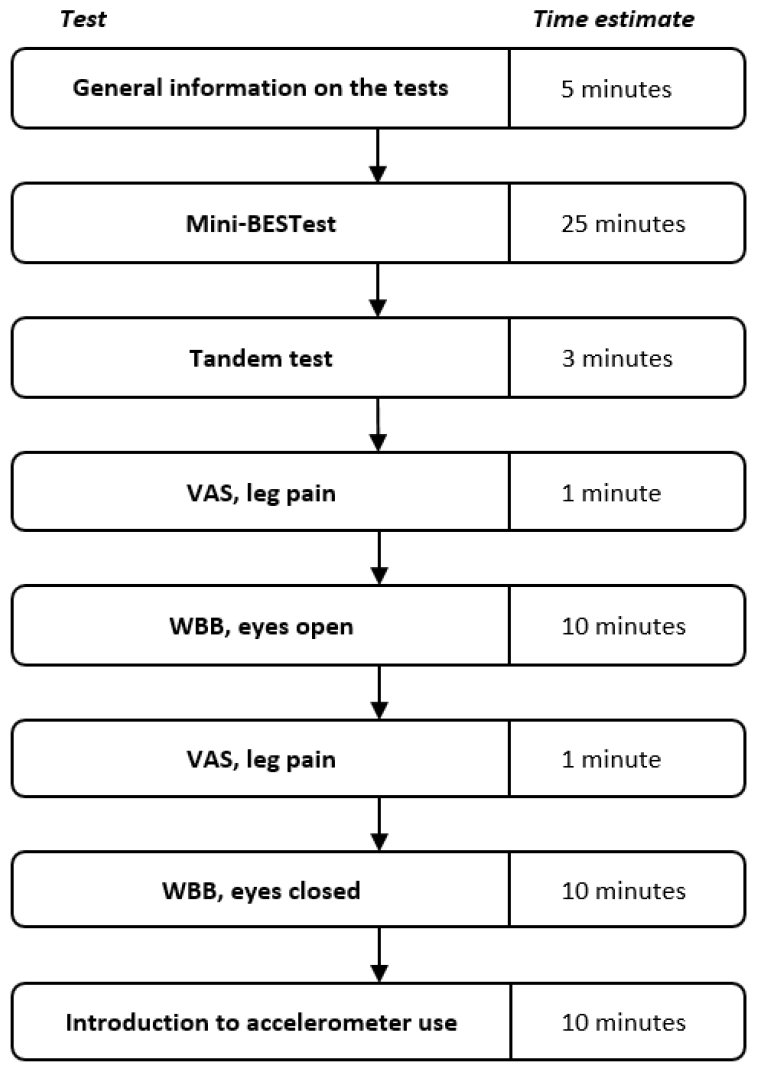
Overview of the order of tests administered during each data collection visit. Total time needed for all tests at each visit is estimated to be 65 minutes. *Mini-BESTest: Mini Balance Evaluation Systems Test, WBB: Wii Balance Board, VAS: Visual Analog Scale*.

### Sample size

Previous studies on the effects of decompressive surgery on postural control and physical activity are sparse and show varying effect sizes depending on methodology and choice of outcome variable. For the calculation of sample size, a conservative effect size of 0.25 was decided upon for both postural control and physical activity measures based on the available literature [21,25,28]. To ensure 80% power with α = 0.05 for a two-sided test, and with an assumption of 10% loss to follow-up, a population of 80 participants will be necessary. An interim analysis of the standard deviation (SD) of the pre-operative data is planned after the first 30 patients have been included, which may result in an amendment to the target sample size if the calculated SD varies greatly from the literature.

### Data collection and analysis

All data pertaining to the study will be entered into a Research Electronic Data Capture (REDCap) database. REDCap is a secure, web-based software platform designed to support data capture for research studies, which enables audit trails for tracking data manipulation and export procedures [66,67]. Full access to the dataset will be limited to the members of the Data Monitoring Committee while a pseudonymized version will be made available to all members of the study group.

Statistical analysis will be conducted using R 4.3.2 (R Core Team, 2021). Patient characteristics and variables of postural control and activity levels at baseline and at each data collection visit will be detailed. Continuous variables will be presented as means with SD or medians with inter-quartile range, depending on normality. Categorical variables will be presented as frequencies and percentages. The Kolmogorov-Smirnov test will be used to test for normality where appropriate.

Differences between means from baseline to 24-months post-operative will be reported with 95% confidence intervals and will be assessed using paired T-tests for continuous variables, Wilcoxon signed-rank test for ordered categorical variables, and McNemar’s test for binary categorical variables.

Multivariable linear regression analysis will be used to adjust for potential confounders and linear regression will be used to test for trend across the data collection visits. Throughout, a p-value of 0.05 will be considered statistically significant.

#### Missing data

We estimate loss to follow-up at 10%. Additionally, data may be missing due to participants missing one or more data collection visits during the study period. Missing data during the post-operative data collection period will be handled using a linear mixed effects model. Patients who do not participate in any post-operative data collection visits will be excluded from analysis.

### Patient and Public Involvement

A Patient Committee (PC) has been assembled during the preliminary study preparations, with the stated purpose of providing patient-directed feedback during the study period. The PC members consist of volunteers from the list of included participants and rotate with respect to availability. Meetings are planned to occur before, during and after the inclusion period, and feedback is gathered concerning, but not limited to, study design, burden of participation, patient recruitment and dissemination of results.

## Data Availability

All data produced in the present study will be available upon reasonable request to the authors.

## ETHICS AND DISSEMINATION

The study will be performed according to the Declaration of Helsinki. The protocol has been approved by the Regional Ethics Committee of Region Zealand (Case ID EMN-2022-08110) and the Danish Data Protection Agency (Case ID REG-100-2022). Major modifications to the study design that necessitate an amendment to the protocol will be sought to be approved by the Regional Ethics Committee of Region Zealand and will be communicated to the participants either electronically or in-person. Participants will be informed of the full scope of the study by licensed health personnel trained by the PI before informed consent is collected, and enrolment in the study will not affect the participants’ future course of treatment. No compensation will be received from study participation. All study-related measurements are non-invasive and are not considered to cause any risk or significant discomfort, and participants are covered by the National Danish Patient Insurance Association in case of any adverse events related to the study.

All results from the study, positive, negative, and inconclusive, will be published in peer-reviewed international journals and presented at national and international scientific meetings. Authorship allocation will follow the Vancouver Recommendations. All participants will receive an invitation to a seminar after the study period has ended, whereupon the results of the study will be presented.

The authors plan to deposit the full dataset at the Danish National Archives after the study period has ended. A pseudonymized version of the dataset will be kept by the authors for an additional 10 years and will be made available to other researchers at reasonable request. The statistical code used during the analysis of the results from the study will be made publicly available.

## AUTHOR CONTRIBUTIONS

The primary design of the study was conceived by RB. OZ, LC, MA, LD and RB developed the final study design, which was implemented across the inclusion centers by OZ, RB and MA. DH and KJ provided statistical expertise and guidance, while the primary statistical analysis will be conducted by OZ, DH and KJ. OZ wrote the protocol in its current form, while DH, KJ, LC, MA, LD and RB contributed to the refinement of the protocol. All authors approved the final manuscript.

